# User perspectives on the Molbio Truenat platform and Tuberculosis assays for decentralized testing in Mozambique and Tanzania

**DOI:** 10.1101/2025.03.16.25324071

**Authors:** Maria del Mar Castro, Celso Khosa, Grace Mhalu, Lonze Ndelwa, Simeon Mwanyonga, Thresphory Zumba, Délio Elísio, Katia Magul, Yolanda Manganhe, Antonio Machiana, Marta Cossa, Raphael Edom, Jerry Hella, Dinis Nguenha, Morten Ruhwald, Vinzeigh Leukes, Adam Penn-Nicholson, Katharina Kranzer, Claudia M. Denkinger, the TB-CAPT Consortium

## Abstract

**Background:** Timely and appropriate diagnosis and treatment are key to end tuberculosis (TB). Incorporating users’ preferences when implementing decentralized strategies for diagnosis may facilitate scale-up and impact. This qualitative study embedded within a cluster randomized controlled trial explored the values and preferences of multiple stakeholders regarding a TB diagnostic strategy using the Truenat platform, MTB Plus and RIF Dx Assays in Mozambique and Tanzania.

**Methods:** We conducted semi-structured interviews with people with presumptive TB (n=34), professional users (laboratory technicians, nurses, clinicians, n=19) and national decision makers (n=5). Direct observations of testing procedures and usability surveys were also conducted. Thematic analysis was performed, informed by the Consolidated Framework for Implementation Research.

**Results:** Facilities varied in testing capacity, number of cases and time-to-results (from same-day to >2 weeks). Availability and supply of reagents and cartridges were described as an issue by healthcare workers, and a potential cause for delayed results. The Truenat platform for detection of TB was considered easy-to-use (median SUS score 90/100) as well as acceptable and fit to the context where the evaluation was conducted. Truenat’s advantage was appreciated in facilities with limited prior testing capacity (e.g., shipping samples, using microscopy), including short time-to-results, reduced need to return to provide more samples and fewer infrastructure needs (compared to GeneXpert). People with presumptive TB preferred the same-day results and rapid initiation of treatment enabled by Truenat testing. Some viewed waiting longer time (>1day) for the results acceptable if it were to result in increased accuracy. Regarding the diagnostic process, participants valued the support and counseling from the healthcare workers.

**Conclusions:** The Truenat platform and TB assays were perceived as easy to use by health providers, and their implementation in decentralized settings was viewed as an acceptable, feasible and preferred alternative to off-site Xpert testing for TB in Mozambique and Tanzania.

## Introduction

Tuberculosis (TB) remains a leading cause of death from a single infectious agent, with an estimated 10 million new cases annually [1]. Despite being preventable and curable, TB disproportionately affects disadvantaged populations [2], with nearly 40% of newly affected individuals remaining undiagnosed. The COVID-19 pandemic exacerbated this issue, causing a rise in TB cases and deaths due to reduced access to diagnostic services [3]. The World Health Organization (WHO) aims to reduce TB deaths by 95% and new cases by 90% by 2035 [4], but achieving these targets requires novel diagnostic tools and interventions to close the significant diagnostic gap.

Molecular diagnostic technologies have marked progress in TB diagnosis and are valued by recipients and providers [5]. Among them, the Xpert MTB/RIF test has been widely implemented after its recommendation by WHO in 2010 [6]. However, its complex infrastructure requirements and centralized deployment have limited its impact [7], since many high-burden countries face challenges such as unreliable power supplies and environmental constraints, that prevent the optimal use of Xpert MTB/RIF in primary health care settings [8, 9]. Consequently, many TB cases remain undiagnosed and untreated, underscoring the need for more accessible and feasible diagnostic solutions. The Molbio Truenat platform (henceforth called Truenat platform), a portable and battery-operated molecular diagnostic tool, offers a promising alternative for decentralized TB diagnosis [10]. Recommended by WHO in 2020, the Truenat system includes the Trueprep DNA extraction device and the Truelab micro-PCR machine, designed for use in peripheral laboratories and primary health clinics with minimal infrastructure. This platform’s portability, simplicity and stability under varied environmental conditions make it more suitable for resource-limited settings, potentially enabling same-day diagnosis and treatment initiation.

Understanding the experiences and preferences of intended users, including patients and healthcare providers, is crucial for the successful implementation and uptake of the Truenat platform. Qualitative methods are ideal for exploring user perspectives, providing deep insights into the contextual and subjective dimensions of diagnostic tool utilization. A qualitative synthesis of studies assessing molecular tests for TB and rifampicin (RIF) resistance testing identified aspects valued by recipients and providers, focusing on Xpert MTB/RIF [5]. Notably, experiences with Molbio Truenat were limited [11]. Thus, assessing the values, preferences, acceptability, and feasibility of implementing the Truenat platform at a primary care facility is essential to ensure the technology meets the needs of its users [12].

This qualitative study sought to investigate user perspectives on the Truenat platform and TB assays of both end-users (people seeking TB diagnosis, henceforth called ‘recipients’) and professional users (laboratory technicians, clinicians, nurses, and decision-makers) in primary health care settings in Tanzania and Mozambique [13]. The study also aimed to understand the feasibility and preferences related to TB diagnosis using the Truenat platform, and to assess its usability and acceptability among the intended users. Through this comprehensive evaluation, we seek to inform policy decisions regarding the implementation and scale-up of the Truenat platform and TB assays in resource-limited settings.

## Methods

### Study Design

This qualitative study was conducted as part of the broader TB-CAPT CORE trial, a pragmatic, cluster-randomized controlled trial aimed at evaluating the effect of implementation of the Truenat platform at primary health clinics for TB diagnosis and treatment initiation within 7 days of enrolment [13].

### Study Settings and Participants

The study was carried out at primary health care clinics in Dar es Salaam and Njombe (Tanzania), as well as Maputo and Manhiça (Mozambique). These sites were selected based on their involvement in the TB-CAPT CORE trial and included both the intervention (onsite molecular testing with Truenat platform and TB assays, process optimization for same-day TB diagnosis and treatment initiation) and control facilities (off-site Xpert testing in all clinics, with parallel on-site smear microscopy in some clinics) [13]. Participants were purposively sampled using a maximum variation approach to ensure diverse perspectives from different stakeholders. Three groups of participants were included: healthcare providers (HCPs), such as nurses, clinicians, laboratory technicians, and study staff involved in the CORE trial, decision makers (DMs - individuals with a direct role in the development of care guidelines, facility management, operational or administrative decision-making in TB programs) and participants (recipients) of the trial.

We sought to include 3-5 healthcare workers per user group (nurses, clinicians, lab-technicians), to reflect the various professional cadres, 10-12 study participants from each coordinating site (20-24 per country who met inclusion criteria of the TB-CAPT CORE trial) and up to 3 DMs per country. All interviewees provided written informed consent and were approached by trained study staff.

### Data Collection Methods

Data were collected through semi-structured interviews, direct observations, and self-administered surveys, as detailed in the study protocol [13]. Local investigators conducted face-to-face semi-structured interviews at the clinics or in locations agreed with the study staff (for DMs). Interviews were conducted privately, with no additional personnel present. A topic guide was developed and pilot-tested with local research teams for each participant group. For recipients, it covered diagnostic processes, sample collection, time-to-results, preferences for TB testing, accessing facilities for testing and follow-ups. The topics for HCPs included samples and challenges related to sampling, testing capacity, results, follow-up, and preferences for testing, while topics for DMs, included the current TB diagnostic landscape, their views on its feasibility of implementation for Truenat and it’s fit into the local contexts. Interviews were conducted in English or the local language and lasted 30 to 60 minutes.

In addition to interviews, trained research staff observed health workers performing specimen collection and testing using the Truenat platform. Observations included recording time spent on tasks, errors or failures, and general comments on device usability. The System Usability Scale (SUS) [14], a 10-item questionnaire on a 5-point Likert scale, was administered to eligible HCPs per country. Additional questions assessed the ease of use and acceptability of the Truenat platform.

### Data Entry and Analysis

After informed consent, all interviews were audio-recorded and complemented with debriefing notes taken by the local investigator. Debriefing sessions [15] between the local investigators and MDMC were conducted via teleconference at the beginning of the study to explore needs of refining the topic guide, challenges and questions from the team. Audio files were transcribed, transcripts were reviewed against audio recordings for accuracy by the local teams and translated to English when necessary. Data were managed using the NVivo software version 14 for qualitative analysis. Thematic analysis [16] was employed to analyze qualitative data, focusing on identifying patterns and themes related to user experiences, challenges, and preferences. Saturation was sought for study participants and healthcare providers, while decision-maker interviews were exploratory. A pre-defined coding framework, considering the constructs of the updated Consolidated Framework for Implementation Research [17], was used and updated as new themes emerged during analysis. Descriptive analysis was conducted for survey data, including calculating SUS scores and summarizing usability ratings. Report follows the COREQ guidelines [18].

### Ethics

The study was conducted in accordance with the Declaration of Helsinki and local regulatory requirements. The trial protocol, including the qualitative part published in this article, was approved by all the following institutional review boards: IHI, NIMR, CISM, INS and the National Bioethics Committee for Health in Mozambique (Ref:217/CNBS/21) and National Health Research and Ethics Committee in Tanzania (Ref: #NIMR/HQ/R.8c/Vol.I/2323) information. The qualitative protocol was also approved by the Ethics Committee of the Heidelberg University Hospital (Ref: S-616/2021)

The study protocol has been published [13] and was registered at ClinicalTrials.gov, NCT04568954. Only adults 18 years of age or older who signed an informed consent were enrolled in the study.

### Patient and public involvement

Patients and public were not involved in the design of this study. However, local health professionals, researchers and organizations were consulted in designing and carrying out study activities and interpreting results.

## Results

We present the perspectives of 59 participants (n=32 from Tanzania, n=27 from Mozambique) involved in the TB-CAPT CORE trial. This includes recipients (individuals with presumptive TB with positive and negative test results), HCPs, and DMs from both intervention and control clinics. Tables 1 and 2 provide a breakdown of participants, while themes are organized according to the updated CFIR framework (Table 3): a) innovation factors, b) inner setting, c) individuals (people receiving healthcare or recipients, HCPs and DMs), and d) outer setting.

**Table 1.** Characteristics of study participants.

| Characteristics | Recipients |  | HCPs and DMs |  |
| --- | --- | --- | --- | --- |
| Country |  |  |  |  |
| Mozambique | 15 | 42.9 | 12 | 50.0 |
| Tanzania | 20 | 57.1 | 12 | 50.0 |
| **Age, years (Median, IQR)** | 36 | (24.5 - 42) | 36 | (30.5 - 43.5) |
| **Sex, n (%)** |  |  |  |  |
| Female | 21 | 60.0 | 15 | 62.5 |
| Male | 14 | 40.0 | 9 | 37.5 |
| **Research coordinating site\*, n (%)** |  |  |  |  |
| CISM | 10 | 28.6 | 6 | 25.0 |
| IHI | 12 | 34.3 | 5 | 20.8 |
| INS | 5 | 14.3 | 6 | 25.0 |
| NIMR | 8 | 22.9 | 7 | 29.2 |
| **TB test result, n (%)** |  |  |  |  |
| Positive | 23 | 65.7 | - | - |
| Negative | 12 | 34.3 | - | - |

\*CISM: Centro de Investigação em Saúde de Manhiça, INS: Instituto Nacional de Saúde, IHI: Ifakara Health Institute, NIMR: National Institute for Medical Research.

**Table 2.** Characteristics of participants in the usability valuation.

| Characteristics | SUS and acceptability<br>(n=12) | Direct observations<br>(n=12) |
| --- | --- | --- |
| **Role** |  |  |
| Laboratory technician or assistant | 9 | 9 |
| Nurse | 3 | 3 |
| **Country** |  |  |
| Mozambique | 4 | 4 |
| Tanzania | 8 | 8 |
| **Working experience** |  |  |
| ≤ 5 years | 2 | 2 |
| 6-10 years | 5 | 5 |
| 10+ years | 5 | 5 |
| **Times device was used** |  |  |
| <3 | 1 | 1 |
| 3-10 | 3 | 3 |
| >10 | 8 | 8 |

**Table 3.** Summary of considerations for implementation of Molbio Truenat in decentralized settings.

| CFIR Domain | Findings |
| --- | --- |
| **Innovation factors** |  |
| Complexity | Trueprep device: difficulties handling error messages |
|  | Use of inadequate volume of buffer |
|  | Error: failure to check for the internal control line |
| Relative advantage | On-site testing and faster results were perceived as a significant improvement |
|  | Reduced infrastructure requirements and the availability of rifampicin resistance testing were appreciated |
|  | Accuracy of Truenat compared to time-intensive microscopy testing |
| Acceptability and perceived fit | Providers and DMs valued the ability to deliver faster results, increase testing capacity and accessibility in rural areas that lack continuous electricity supply |
|  | People under evaluation for TB appreciated the same-day results and treatment initiation with Truenat |
|  | HCPs stated willingness to adopt the technology more broadly |
| **Inner setting** |  |
| Feasibility and integration in facility workflow | Molecular testing on site was perceived as aligned with the role of the HCPs and that benefits of on-site testing outweighed the additional efforts and changes in the work schedule |
|  | HCPs valued the increased efficiency with Truenat. Taking only one spot sample, also reduced the number of tasks HCPs needed to do |
|  | Testing capacity is determined by the throughput of available devices |
| Local infrastructure | Requirement of adequate spaces for handling samples (procurement, reception and processing) and a fixed place to locate the device |
|  | HCPs expressed concerns about the risk of infection |
| **Individuals** |  |
| Preferences | Preference for decentralized testing to minimize costs, improve access, and provide faster diagnosis and treatment for tuberculosis |
|  | Most people preferred same-day results to reduce economic burden (transportation and loss of work), fears and stress while waiting to know their condition |
| Recipient centeredness | Recipients valued the support and counseling provided by healthcare staff during the diagnostic process and treatment |
| **Context / outer setting** |  |
| One size does not fit all | The choice of technology should be guided by the specific needs and context |
| Supply of reagents and cartridges | Limited availability of reagents and cartridges was a recurring concern across both countries |
|  | Adding Truenat to the repertoire of molecular TB testing technologies helps reduce reliance on a single provider |
| Infrastructure and costs | Truenat's ability to operate without skilled personnel or a continuous power supply was seen as ideal for decentralized settings |
|  | Need for budgeting, training, and infrastructure improvements to ensure the successful implementation of Truenat |

### Innovation factors

#### Complexity: usability and ease-of-use

User experiences with the Truenat platform were largely positive, with a System Usability Scale (SUS) mean score of 90 (range=65-92.5). All 12 surveyed users (table 2) reported confidence in operating the instrument, and most found components like the Trueprep DNA extraction device and Truelab micro-PCR machine easy to use. The most common error was failure to check for the internal control line of the Trueprep cartridge prior to use, in 50% of observations (n=6/12). Interviewees consistently described Truenat as user-friendly, however, challenges with the Trueprep device, such as buffer volume errors and reprocessing of samples, underscored the need for ongoing training.

*“First, I’ve noticed that the machine is as friendly as it is. […] I can do it because it is a machine that is friendly to me.” (HCP, Tanzania)*

*“[My] difficulties found in using Trueprep were errors related to the amount of buffer for the amount of sample”. (HCP, Mozambique)*

Some participants highlighted difficulties with handling errors from the Trueprep device and emphasized the need for continued support to address these issues effectively.

#### Relative advantage

Participants described Truenat’s relative advantage over existing diagnostic methods, namely off site Xpert MTB/RIF testing. In these settings, Truenat’s ability to deliver same day results was a significant improvement over traditional microscopy or the need to transport samples for molecular testing. Facilities for the trial were selected based on the absence of on-site molecular testing; however, some HCPs had experienced using GeneXpert for routine care, enabling comparisons between the two technologies. Reduced infrastructure requirements compared to GeneXpert, and the availability of rifampicin resistance testing (compared to microscopy) were seen as key advantages of Truenat. In addition, some providers highlighted the accuracy of Truenat compared to time-intensive microscopy testing and described trusting the evidence supporting its use.

*“It is a good machine, it brings correct results, and it has been researched properly” (HCP, Tanzania)*

*“I consider it effective, because, well, it’s a platform where the result is seen on the same day. It doesn’t take long, we’re talking about an hour and 30 minutes, right? So, I think it’s effective.” (HCP, Mozambique)*

#### Acceptability and perceived fit

Truenat was perceived as acceptable and compatible with the needs of healthcare facilities where it was evaluated. Users valued the ability to deliver faster results, increase testing capacity and accessibility in rural areas that lack continuous electricity supply (“*Unlike GeneXpert which requires electricity, this can be used even in villages where there is no electricity, and it can provide answers quickly*” - HCP, Tanzania). The implementation of Truenat was seen as a significant step towards eradicating TB due to its possible impact on case detection across various healthcare settings, potentially reducing transmission.

*“Thank you for Truenat, because it really helps us a lot. In the past, saying a patient comes back in 4 days was terrible”.* (HCP, Mozambique)

*“Personally, I really like the Truenat device because it makes it easier for us to serve our clients. It is different from the microscope we used to use which had many processes before giving results. […] With the microscope, we would take a long time, and a client could give their sputum and wait for two weeks before getting results. In case they were positive, it would mean that they had already spread the disease to others. But with this device, we can give results to the patient within 24 hours, making it easier to give them treatment and prevent them from infecting others.” (HCP, Tanzania)*

People diagnosed using Truenat appreciated the same-day results and treatment initiation; some compared the current service with previous personal experiences or from relatives, when long time to results were common.

*“My people back at home, […] they were once sick and their test results took three days so, when I heard that, I thought it’s going to be the same with me and it will take two to three days, but with this [Truenat] it’s easy. […] I was happy with that because you know if you are sick and you got the results of what is making you sick then there is a light already, I have tested and I have started my medications.” (Recipient, positive TB test, intervention arm. Tanzania)*

The same-day diagnosis facilitated quicker clinical decisions, resulting in better recipient care and willingness from providers to adopt the technology more broadly. HCPs expressed a desire for the government to endorse wider implementation of Truenat, recognizing its potential for improving case detection in decentralized settings.

“*I really wish that the government can approve this device so that it can be used in the hospital*.” (HCP, Tanzania)

### Inner setting

#### Integrating Truenat into facility workflows: feasibility and operational efficiency of molecular testing

Facilities varied in terms of testing capacity and daily number of presumptive cases identified. Molecular testing on site was perceived as fitting the workflow at the facility and aligned with the role of the HCPs, who perceived that the benefits of on-site testing outweighed the additional efforts and changes in the work schedule. In facilities that did not have prior testing capacity, and in control facilities, HCPs described how the lack of local testing increased the risk of attrition and required follow-up activities, like additional clinical encounters or home visits.

*“Actually, I think the challenge we have is the lack of an internal laboratory. This makes the diagnosis part difficult; it makes it very difficult. There are cases that we end up losing because patients are impatient […] As long as we don’t have an internal laboratory to work, we will have many problems to follow TB patients”. (HCP, Mozambique)*

Changes to the workflow with Truenat included, in addition to running the test, the documentation of results and notification. When compared to microscopy, Truenat increased efficiency and the number of cases tested per day. Taking only one spot sample, also reduced the number of tasks HCPs needed to do.

*“Truenat of course changed the schedule, so changing it, the patient is coming today, and I will treat him and finish with him, if I need to start treatment, I will start it. But if the old device is used, the patient will come today, and you will have to wait for him tomorrow, so it takes you a long time to serve him and also wastes his time.” (HCP, Tanzania)*

One HCP indicated that the on-site testing also improved the reporting of cases from the facility.

*“It has helped me because I have to send reports weekly of how many have we managed to test, and how many suspected cases have been raised, and if it was not for this tool, it could be hard and that is the reason (name) health center has new TB cases” (HCP, Tanzania)*

However, the testing capacity is determined by the throughput of available testing devices. In facilities with a high volume of samples for evaluation each day, the number of tests that can be run simultaneously is a factor that affects the timely delivery of results. In addition, despite improvements for diagnosis of new cases, challenges persist for treatment monitoring where microscopy continues to be used.

#### Infrastructure gaps and infection control: facility improvements needed for safe Truenat use

Despite the low infrastructure requirements of Truenat, HCPs and DMs highlighted the need of improving the infrastructure at the facility, including adequate spaces for handling samples (procurement, reception and processing) and dedicated working surface for the device. HCPs from both Mozambique and Tanzania expressed concerns about infection control and the risk of transmission given the limited infrastructure.

*“I am currently processing samples outdoors and that is not safe for patients at that time […] and not only for patients, for health unit professionals, it is not safe at all.” (HCP, Mozambique)*.

### Individuals

#### Decentralized testing as a preferred model: enhancing convenience and quality of care

Overall, recipients and HCPs preferred decentralized testing. For HCPs, the ability to process the samples locally was highly valued, while transportation of samples to centralized facilities for testing was less preferred due to inconvenience and delays in obtaining results. Some HCPs mentioned that loss of samples, sample spillage and delays receiving the results were risks associated with sample transport to off-site facilities. People under evaluation for TB mostly preferred facility-based testing, ideally at the nearest facility or one that offers diagnostic services. This was driven by aspects related to quality of care, and the perception that their issues can be resolved at the facility. For those preferring home or community-based testing, convenience was the most cited reason.

*“Maybe the health unit could make it easier for people who have difficulties to get around, yes, sent its brigades to these people to be diagnosed at home, because […] for people to come to the health unit, it only depends on them, so it gets a little complicated. (Recipient, positive TB test, control arm. Mozambique)*

HCPs reported positive experiences with on-site Truenat testing, including the capability to offer improved care, enhance surveillance, and feeling empowered to take clinical decisions in a timely manner.

“You can *test a patient even when there is no electricity, instead of telling a patient to go and come back when there is electricity. Again, it has helped in the testing and treating issue; like you test and start the medications on the same day, different from when we were testing and asking patients to come the next day for the result. So, [it] has generally given us the morale as even our HIV patients can be tested and start their treatments on the same day.”* (HCP, Tanzania)

#### Balancing accuracy and convenience: preferences for sample type and time-to-results

Accuracy was an important driver of preferences and was often cited as a trade-off for other features of diagnostic services. In general, people under evaluation for TB valued test accuracy higher than other factors like location of testing, type of sample or time-to-results.

Within the study, time to results varied from receiving results on the same day of submitting the sample versus two weeks. Facilities who had to transport samples to off-site laboratories generally experience longer delays in receiving the results. Recipients in the control facilities often described the need for multiple visits, due to reasons like providing ‘morning’ samples, additional testing or collecting results. They were stressed and worried about their condition, and often decided to self-medicate. HCPs also describe prescribing antibiotics while waiting for test results. Among those receiving same-day results, waiting times at the facility were variable (1-5h). Depending on the distance to the facility, some participants went home and returned to collect the results later in the day, while those waiting at the facility expressed discomfort and boredom during the wait. Overall, the preference was for faster diagnosis.

*“Frankly I felt bad and based on the condition I was I wanted to be well, so I felt like it was a lot [of time], I was coughing a lot, and I had to wait for the next three days. I did not just wait without any action, so I took some coughing and fever medicines to help me while waiting for the results because my condition was bad.” (Recipient, negative TB test. Tanzania)*

In terms of samples, HCPs highlighted the challenges of obtaining any or good quality sputum samples especially from children and people living with HIV and acknowledged other diagnostic modalities like chest X-rays (CXR). However, among recipients, sputum was the specimen of first choice, due to familiarity with the sample, as many of them had previously been investigated for TB or they knew a person (relative, neighbor) who had been diagnosed with TB; they also recognize that the disease affects the lungs and hence it was “logical” to investigate a respiratory sample.

*“I cannot say there is another one [sample] that I believe and trust because I do not know any other, what I know is the sputum only.*” *(Recipient, negative TB test, intervention arm. Tanzania)*

Some recipients described blood and urine as more convenient samples. Two respondents mentioned preferring stool. In addition, although not a sample, multiple participants mention X-Rays as an acceptable alternative to know their condition, in part due to the possibility to identify alternative diagnostics besides TB.

*“Now it’s easier to urinate. Because now I no longer have a cough […] now I don’t have sputum. Even blood is easy for me.” (Recipient, positive TB test, intervention arm. Mozambique)*

#### Recipient centeredness: value of compassionate care and counseling

Recipients valued the support and counseling provided by HCPs during the diagnostic process and treatment. However, fears of stigmatization following a TB diagnosis were prevalent, underscoring the need for ongoing support and education to address these concerns.

*“I have had many challenges in getting this checkup in any way in any place and I have never gotten results, however when I used this instrument, I was very grateful and the health care providers from this place are very cooperating with good outstanding manners they don’t look down on us. As they follow up to advise and now, we get quick results, and we work nicely with the health care provider […]” (Recipient, positive TB test, intervention arm. Tanzania)*

HCPs described multiple strategies to tailor service delivery, such as prioritizing individuals living farther from the facility, conducting outreach to vulnerable groups, and adapting communication to match the recipients’ literacy levels and understanding.

*“We once got one from [village], so I had to talk to the lab technician, because he came from afar, we asked him to wait for two hours, and the technician gave him priority.” (HCP, Tanzania)*.

### Context / outer setting

*One size does not fit all: program goals for molecular testing, and feasibility of implementation*.

DMs highlighted the role of international organizations and donors in procuring equipment and supplies for the implementation of molecular TB diagnostics. Endorsement by WHO of the molecular diagnostic technologies was an important factor in the decision to adopt them. Respondents indicate that GeneXpert and microscopy were the most widely used diagnostic tools. The addition technologies like Truenat (and urine LAM) presents opportunities for decentralized testing, in line with the program goals of strengthening diagnostic services and increasing coverage.

Overall, there was a consensus on the importance of widespread access to modern diagnostic tools for TB management. Recipients and HCPs expressed preference for decentralized testing to minimize costs, improve access, and provide faster diagnosis and treatment for TB. Truenat’s ability to operate without a continuous power supply was seen as ideal for these settings. However, DMs stressed that “one size does not fit all,” and the choice of technology should be guided by the specific needs and context, rather than adopting a single solution.

*”[…] so that we can advise the ministry to scale up a certain machine more than another machine or one technology than another, […] as I said at the beginning, one size does not fit to all, you can say [a] technology cannot go all over Tanzania, it can be suitable for some zones and the other one can fit another part differently” (DM, Tanzania)*

#### Supply disruptions: Importance of multiple providers and diagnostic technologies

Limited availability of reagents and cartridges was a recurring concern across both countries. Interviewees described instances of international suppliers splitting deliveries into smaller batches due to limited availability, as well as supply gaps affecting their ability to meet testing targets. At the facility level, HCPs also describe issues with the availability of supplies and reagents for all types of testing, including microscopy, GeneXpert and Truenat.

*“I think it’s very good, because if we always had the machine, it would improve our work, but I would ask that we never run out of cartridges because in the general diagnosis of TB we always have cartridge problems.” (HCP, Mozambique)*.

One DM emphasized the positive impact of adding Truenat to the repertoire of molecular TB testing technologies, noting that it helps reduce reliance on a single provider and breaks the “monopoly” in TB molecular diagnostics.

“*As I previously stated, the issue with GeneXpert is obtaining cartridges because the company that manufactures it cannot meet the demand, so we can rely on Truenat to alleviate these difficulties.” (DM, Tanzania)*

#### Costs, training, and cross-program collaboration for Truenat implementation

DMs discussed the need for budgeting, training, and infrastructure improvements to ensure the successful implementation of Truenat. Strengthening collaboration between TB and HIV programs, increasing treatment coverage, and improving diagnostic services were also highlighted as essential strategies for sustainable TB management. The involvement of international organizations and external donors to provide equipment and resources for Truenat’s implementation was described.

*“The main challenge in any strategic plan may be establishing an adequate budget for activity implementation. When you want to train a health care provider, for example, you end up with untargeted plans.” (DM, Tanzania)*

#### Impact on equity and access

Truenat was felt to reduce the number of visits to the facility, potentially reducing the costs to people with suspected TB, including transportation and lodging. Although testing is generally free of cost and some recipients do not spend money in transportation (e.g., travel by foot) the time spent in the clinic, and the additional visits, were perceived as burdensome.

*“I didn’t like spending my money on all these trips. It turned out to be a time-consuming process and for a sick person it is not easy.” (Recipient, negative TB test, control arm. Mozambique)*

## Discussion

Our study presents the perspectives of multiple type of users and decision makers for Truenat implementation. Overall, the platform and TB assays demonstrated high usability and acceptability among recipients, HCPs and DMs. The ability to provide same-day results and the option to run the device without a continuous electricity supply were seen as advantageous for its use in decentralized settings and contributed to the perceived fit to the evaluated settings. Despite the low infrastructure requirements, users described challenges related to supply chain management, throughput, and local infrastructure, such as dedicated bench space for sample processing. These infrastructure improvements are necessary to ensure safe and effective use, particularly for healthcare providers, who face a high risk of workplace infection [19]. The need for ongoing training and support was also identified.

In terms of usability, our findings align with a previous systematic review on molecular TB testing [5], a recent study among operators of Truenat in Nigeria [11] and reports of operational research for Truenat implementation by the STOP TB partnership [20]. The most frequently reported issues were related to sample processing with Trueprep, similar to previous operational studies [11, 21–23]. Although not widely discussed in the interviews, issues with invalid and a high proportion of indeterminate results were identified at trial sites. Interviewees mostly discussed the additional time and effort required to reprocess samples. To overcome this challenge, continued training, support and champions (superusers) to mentor local healthcare providers are recommended, along with continuous site monitoring and retraining as needed to reduce error rates and unsuccessful results [23].

In terms of recipient preferences, sputum remained the preferred sample, despite common descriptions of challenges to produce, handle and transport it. HCPs also mentioned issues with sputum quality, as reported in other studies [5]. Differences in the perceived trustworthiness of non-sputum samples have been reported for urine LAM, stemming from how people associate pulmonary TB and coughing, with sputum [24]. The long history and familiarity of sputum-based testing may also explain its preference, as seen in a multi-country Discrete Choice Experiment regarding TB testing [25]. However, the use of alternative samples may help overcome barriers related to sputum quality and risk of infection [26].

Supply chain challenges and reagent shortages, similar to those seen with GeneXpert, could also hinder Truenat’s implementation [27]. Both providers and decision-makers emphasized the importance of securing a reliable supply chain to meet program goals, thus, tailored procurement strategies are important for implementing Truenat [20]. Engaging multiple suppliers could lower costs and enhance supply sustainability by fostering competition [28]. Expanding the range of available diagnostic tools, including Truenat and other novel tests, would help achieve this. Understanding the local testing needs may also guide the type of device most suitable to the context, since the throughput of the molecular test platforms like GeneXpert and Truenat may affect the workflow at the facility and the ability to offer same-day results. This has been described previously for molecular and other tests like urine LAM [5, 29].

This study has several strengths. First, it benefits from a multi-country evaluation, providing insights into the implementation of Truenat in diverse healthcare settings across Tanzania and Mozambique. The involvement of local investigators, who are knowledgeable of the local languages and context, ensured that the interviews were culturally appropriate and well-understood by participants. Furthermore, the use of standard topic guides and regular debriefings [15] helped maintain consistency across sites and improve the reliability of the data. The analysis was guided by the updated CFIR framework [17], a widely known and utilized tool for understanding the determinants of implementation, enhancing the rigor and depth of the findings and helping to identify key factors that influence the successful integration of Truenat into existing health systems.

This study also has some limitations. The timing of the interviews, conducted during the early stages of implementation, may have limited our ability to capture later-stage challenges that could emerge as the program matures. Additionally, the topic guide did not include aspects such as waste management, the impact of negative test results (not being diagnosed with TB) on recipients and clinical decision making, which could have provided further insights into the operational environment. Participant feedback on the findings was not collected; however, findings were discussed with local investigators and presented to local audiences as part of the study dissemination efforts. Finally, the study did not include children, a population that faces unique diagnostic challenges, such as difficulty producing sputum, and may require further research.

## Conclusion

The Truenat platform demonstrated high usability, feasibility, and acceptability among both healthcare providers and people seeking TB diagnosis. The ability to provide same-day results and operate without a continuous electricity supply were significant advantages, particularly in resource-limited settings. Ensuring adequate supply chain management, improvements in physical infrastructure for sample processing, along with ongoing training and support are needed for sustainable implementation of Truenat. These findings will inform policy decisions and strategies for the broader implementation and scale-up of the Truenat platform in similar settings.

## Supporting information

Supplemental file - TB-CAPT Consortium

## Data Availability

All data produced in the present study are available upon reasonable request to the authors.

## Acknowledgements

We thank the people undergoing TB testing, healthcare providers and decision makers who participated in this study, as well as all members of the TB-CAPT Consortium. We also extended our thanks to Molbio (Goa, India) for donating Truenat platforms and MTB-Plus and MTB-RIF Dx kits for the TB-CAPT trial, where this study is nested, and to the Foundation for Innovative New Diagnostics for coordinating the donation.

