## Supplemental file - TB-CAPT Consortium for "User perspectives on the Molbio Truenat platform and Tuberculosis assays for decentralized testing in Mozambique and Tanzania"

Vinzeigh Leukes[1], Adam Penn-Nicholson[1], Morten Ruhwald[1], Berra Erkosar[1], Mikaela Watson[1], Samuel G. Schumacher[1], Sunita Singh[1], Bernard Kivuma[2], Muhuminu Nuru[2], Francisca Chuwa[2], Omary Ngome[2], Neema Shija[2], Deogratias Bulime[2], Dorcas Mnzava[2], Petro Sabuni[2], Hosiana Temba[2], Jamali Siru[2], Jerry Hella[2], Jonathan Msafiri[2], Maja Weisser[2,16,23,24], Mohamed Mbaruku[2], Mohamed Sasamalo[2], Alice Leonard[2], Ambilikile Malango[2], Annastazia Alexander[2], Faith Komakoma[2], Gloria Msigala[2], Kasmir Johaness[2], Grace Mhalu[2], Robert Ndege[216,23], Swalehe Masoud[2], Theonestina Byakuzana[2,], Mahmud Mahmud[2], Lewis Batao[2], Frederick Haraka[2], Anange Lwilla[3] Craysophy Zachariah[3], Chacha Mangu[3], Emmanuel Sichone[3], Lonze Ndelwa[3], Sara Kiula[3], Alfred Danda[3], Regino Mgaya[3], Bariki Mtafya[3], Issa Sabi[3], Last Mwaipopo[3], Nyanda Elias Ntinginya[3], Raphael Edom[3], Willyhelmina Olomi[3], Delio Elisio[4], Dinis Nguenha[4],[22], Edson Mambuque[4], Joaquim Cossa[4], Marta Cossa[4], Neide Gomes[4], Patricia Manjate[4], Shilzia Munguambe[4], Sozinho Acacio[4], Belen Saavedra[4], Helio Chiconela[4], Katia Ribeiro[4], António Machiana[5], Bindiya Meggi[5], Candido Azize Junior[5], Carla Madeira[5], Celso Khosa[5], Claudio Bila[5], Denise Floripes[5], Ezequiel Nhamtumbo[5], Jorge Ribeiro[5] Sofia Viegas[5], Albero Garcia-Basteiro[4,6], Belén Saavedra[6], Carole Amroune[6], Joanna Ehrlich[6], Laura de la Torre Pérez[6], Sergi Sanz[6,25,26], Friedrich Riess[7,20], Katharina Kranzer[7,12,,20], Michael Hoelscher[7,18,19,20], Norbert Heinrich[7,18,19,20], Sarah Mutuku[7,20], Tejaswi Appalarowthu[7,20], Leyla Larsson[7,20], Maria del Mar Castro Noriega[8,21], Claudia M. Denkinger[1,8,21], Saima Bashir[8], Daniela Maria Cirillo[9], Elisa Tagliani[9], Federico Di Marco[9], Virginia Batignani[9], Akash Malhotra[10, 29], David Dowdy[10], Claudia Schacht[11], Julia Buech[11], Caroline Stöhr[11], Marguerite Massinga Loembé[13], Pascale Ondoa[13], Nqobile Ndlovu[13], Fumbani Brown[13], Yonas Ghebrekristos[14], Cindy Hayes[14], Ilse vanderwalt[14], Shareef Abrahams[14], Puleng Marokane[14], Mbuti Radebe[14], Neil Martinson[14], Anura David[15],Lesley Scott[15], Pedro Da Silva[15], Riffat Munir[15], Wendy Stevens[14, 15], Charles Abongomera[16,23], Klaus Reither[16,23], Leon Stieger[16, 23], Adrian Brink[17], Chad Centner[17], Helen Cox[17], Judi van Heerden[17], Mark Nicol[27], Nchimunya Hapeela[17], Parveen Brown[17], Reyhana Solomon[17], Widaad Zemanay[17], Tania Dolby[28].

[1]FIND, Geneva, Switzerland

[2]Ifakara Health Institute, Dar es Salaam, Tanzania

[3]Mbeya Medical Research Centre, National Institute for Medical Research (NIMR), Mbeya, Tanzania

[4]Centro de Investigação em Saúde de Manhiça (CISM) Manhica, Mozambique

[5]Instituto Nacional de Saúde (INS), Marracuene, Mozambique

[6]ISGlobal, Hospital Clínic – Universitat de Barcelona, Barcelona, Spain.

[7]Institute of Infectious Diseases and Tropical Medicine, LMU University Hospital, LMU Munich, Germany

[8]Department of Infectious Disease and Tropical Medicine, Heidelberg University Hospital, Heidelberg, Germany

[9]Emerging Bacterial Pathogens Unit, IRCCS San Raffaele Scientific Institute, Milan, Italy

[10]Johns Hopkins University Bloomberg School of Public Health, Baltimore, Maryland, USA

[11]LINQ Management GmbH, Berlin, Germany

[12]Clinical Research Department, London School of Hygiene and Tropical Medicine, London, UK.

[13]African Society for Laboratory Medicine, Addis Ababa, Ethiopia

[14]National Health Laboratory Service, South Africa

[15]WITS Diagnostic Innovation Hub, Faculty of Health sciences, University of the Witwatersrand, Johannesburg, South Africa

[16]Swiss Tropical and Public Health Institute, Allschwil, Switzerland

[17]Division of Medical Microbiology, University of Cape Town, South Africa

[18]Fraunhofer Institute for Translational Medicine and Pharmacology ITMP; Immunology, Infection and Pandemic Research, Munich, Germany

[19]Unit Global Health, Helmholtz Zentrum München, German Research Center for Environmental Health (HMGU), Neuherberg, Germany

[20]German Center for Infection Research (DZIF), Munich Partner Site, Munich, Germany

[21]German Center for Infection Research (DZIF), Heidelberg Partner Site, Heidelberg, Germany

[22]Department of Global Health and Amsterdam Institute for Global Health and Development, Amsterdam University Medical Centers Location University of Amsterdam, Amsterdam, Netherlands

[23]University of Basel, Basel, Switzerland

[24]Division of Infectious Diseases, University Hospital Basel, Basel, Switzerland

[25]Department of Basic Clinical Practice, Faculty of Medicine, University of Barcelona, 08007 Barcelona, Spain.

[26]CIBER Epidemiología y Salud Pública (CIBERESP), Instituto de Salud Carlos III, 28029 Madrid, Spain

[27]University of Western Australia, Australia

[28]National Health Laboratory Service, South Africa

[29]University of Washington, Seattle, USA
